# A Safety and Feasibility Trial Protocol of Metformin in Infants after Perinatal Brain Injury

**DOI:** 10.1101/2024.05.26.24307924

**Authors:** A Hutchinson, R Pais, AN Edginton, B Pilon, JM MacDonald, ME MacDonald, T Lewis, M Offringa, BT Kalish

## Abstract

**Introduction:** Infants with hypoxic-ischemic encephalopathy (HIE) are at high risk for neurodevelopmental impairment, despite current standards of care. Adjunctive treatments to promote brain repair are needed. The antidiabetic drug metformin has recently been recognized as a neurorestorative agent, but, to date, has not been used in infants. Herein, we describe a clinical trial with the aim of demonstrating the safety and feasibility of metformin use to improve neurodevelopmental outcomes in infants with HIE.

**Methods and Analysis:** In collaboration with patient and family stakeholders, we designed a pragmatic clinical trial to assess the safety and feasibility of metformin administration in infants. To determine appropriate dosing of metformin, we performed Physiologically Based Pharmaco-Kinetic (PBPK) modeling after scaling a published adult PBPK model of metformin to an infant population of full-term newborns to 3-month-olds. Based on this PBPK modeling and target drug exposure, we determined an optimal target dose of 25 mg/kg/day. Trial participants will complete baseline bloodwork and then receive 6 weeks of daily metformin at 50% of the target dose (12.5mg/kg). At a mid-study visit, repeat laboratory testing will be done, followed by an additional 6 weeks of metformin at target dosing of 25mg/kg. The final study visit will include repeat labs following therapy at target dosing. Pharmacokinetics of metformin will be evaluated with bloodwork collected at study visits, as well as an optional at-home monitoring sub-study. The incidence of safety events and feasibility measures will be reported using descriptive statistics. Our infant PBPK model will be validated with study samples and the dose for future trials adjusted based on new knowledge about metformin PK in infants.

**Ethics and Dissemination:** Approval of the Boston Children’s Hospital Research Ethics Committee will be obtained prior to study initiation. Trial oversight will be under the direction of a Data Safety Monitoring Board (DSMB) composed of individuals with the appropriate expertise, including external neonatologists and pharmacologists.

**Registration:** This study has been registered at www.clinicaltrials.gov under NCT06429007.

**WHAT IS ALREADY KNOWN ON THIS TOPIC:**

- Hypoxic-ischemic encephalopathy (HIE) is a major cause of newborn death and neurodevelopmental morbidity.
- Therapeutic hypothermia reduces the risk of death or disability in infants with HIE, but nonetheless, a large proportion of infants experience long term sequelae.
- Metformin is an anti-diabetic drug that has shown promise to promote recovery after brain injury, but it has not been widely used in infants or young children.

**WHAT THIS STUDY HOPES TO ADD:**

- To investigate whether the administration of metformin in infants with a history of HIE is safe and feasible.
- To investigate the pharmacokinetics of metformin in infants.

**HOW THIS STUDY MIGHT AFFECT RESEARCH, PRACTICE, OR POLICY:**

- The results of this trial will serve as the foundation for future clinical trials to study the efficacy of metformin in improving neurodevelopmental outcomes in children with HIE.

## INTRODUCTION

Hypoxic-ischemic encephalopathy (HIE) is caused by reduction in oxygen-rich blood delivery to the brain during the perinatal period. Despite advances in perinatal care, moderate-to-severe acute perinatal HIE in late preterm and term infants remains a critical cause of mortality, severe neurological injury, and subsequent long-term neurodevelopmental disability.^1^ Therapeutic hypothermia (TH) is the current standard of care for neonates presenting with moderate to severe HIE and has been shown to reduce mortality and improve neurodevelopmental outcomes.^2^ However, despite TH, approximately 40-50% of infants with moderate-severe HIE will die or suffer significant neurodevelopmental morbidity.^3^ There is great need to identify and evaluate novel, adjunctive therapeutics for HIE to further protect newborns against the sequalae of early life brain injury. To date, most attention has focused on developing neuroprotective adjuncts that target the secondary phase of brain injury after HIE, during which there is bioenergetic failure.^4^ Unfortunately, despite many years of evaluative research, no safe and effective therapy has reached the bedside.

In recent years, the drug metformin has been recognized as a potential neuroprotective agent, with numerous preclinical and clinical studies demonstrating positive impacts on neurogenesis and neuroplasticity.^5–9^ Metformin has been used as an anti-hyperglycaemic drug for over 60 years with very limited safety concerns.^10^ However, metformin has not been administered to infants after perinatal brain injury. Although limited, there is evidence in paediatric populations to support the neuroprotective and neurorestorative features of metformin. A 12-week pilot trial in 24 children aged 2 to 13 years experiencing cognitive impairment due to cranial radiation showed that children on metformin yielded better performance on tests of declarative and working memory.^9^ Given the unmet needs of patients with HIE, there is a need to evaluate the feasibility and safety of metformin administration in infants. One of the challenges in this population is to determine the correct dose, frequency and duration of metformin that will yield efficacy without causing adverse effects. We designed a clinical trial to evaluate the safety and feasibility of metformin treatment in infants after perinatal brain injury. This manuscript reports the results of *in silico* dose determination and the protocol for a safety and feasibility study that was co-designed by patient and family advocates.

## OBJECTIVE

The objectives of this study are to determine (1) Feasibility and safety of metformin administration in term and near-term infants with HIE; (2) Satisfaction of key stakeholders (parents/caregivers) with the intervention; (3) Validity of our pharmacokinetic model of metformin and to optimize dose for future efficacy-targeted clinical trials. We hypothesize that the short-term administration of metformin to term infants with HIE will be feasible and safe, and that parents and caregivers will be satisfied with this intervention.

## METHODS AND ANALYSIS

This protocol for a pragmatic safety and feasibility trial of metformin use in infants was co-developed in collaboration with families of infants who have sustained HIE and not-for-profit stakeholders to ensure a patient and family-centered study design.

### (A) Patient Population

The study will aim to recruit 30 infants. The inclusion criteria for this study will be infants born > 35 weeks’ gestational age with a clinical diagnosis of HIE, who receive therapeutic hypothermia. Infants will be excluded from study participation in the event of a known genetic or chromosomal disorder, and in the presence of congenital or acquired liver or kidney disease that might, in the opinion of the Principal Investigator (PI) or delegate, affect drug metabolism. Infants will also be excluded if there is maternal use of metformin while actively breastfeeding, if infant weight is below the 10^th^ percentile based on WHO growth charts at the time of study drug initiation, and in the event of any condition or diagnosis, that could in the opinion of the PI or delegate, interfere with the participant’s ability to comply with study instructions, might confound the interpretation of the study results, or put the participant at risk.

Infants will initiate the study intervention between 3 and 6 months of age. Term infants with HIE are hospitalized for a brief period in the inpatient setting, and therefore, this study is designed as a pragmatic outpatient interventional study. This design is necessitated by the fact that the patient population of interest (young children with a history HIE) is almost uniformly at home during the broad critical period of neuroplasticity and potential intervention window. These patients are not hospitalized, and hospitalization would bear a significant practical burden to families. The acute neonatal period most proximal to the time of brain injury is characterized by multi-organ dysfunction, which poses significant risks and pharmacologic considerations for novel neuroprotective drug therapy. Therefore, the investigators feel that an intervention relatively distant from the time of acute organ insult (e.g. 3 months) would be safest and most practical.

### (B) Intervention

Infants are a unique population for drug dosing due to rapid changes in body composition, expression and function of metabolic liver enzymes, and renal drug clearance. These factors lead to, compared to older children and adults, different behaviors of drug absorption, distribution, metabolism, and excretion. The selection of the appropriate drug dose is a critical component of a successful neonatal trial, as under-dosing could lead to insufficient drug-to-target exposure and lack of efficacy, while over-dosing could lead to toxicity.^11^ Therefore, members of our study team established a first-of-its kind neonatal model of metformin pharmacokinetics to facilitate the design of a drug dosing scheme that will maximize the likelihood of efficacy while minimizing the risk of toxicity. We performed Physiologically Based Pharmacokinetic (PBPK) modeling to identify an appropriate starting dose and scaled a published and qualified adult PBPK model of metformin to an infant population of 3-month-olds, with subsequent use for dose-determination.^12^ Virtual children were created using appropriate anatomy and physiology and, particularly with metformin for which MATE1, PMAT, OCT1 and OCT2 play a key role in clearance, the ontogeny of these transporters was incorporated.^12^ To evaluate the ability of the scaled adult model to predict pediatric pharmacokinetics (PK), a clinical trial in 9-year-olds for whom PK was assessed empirically was recapitulated^13^; the mean (n=4) observed plasma concentration time profiles was within the 90^th^ prediction interval. To establish the target dose for the proposed study, the steady state area under the curve (AUC) following daily oral metformin dosing (1000 mg/m^2^ metformin-HCL) in a population of 8–20-year-olds for whom metformin was found to be efficacious for neuroprotection, was simulated.^9^ This AUC was used to derive a weight-normalized metformin dose, in the fed and fasted states, that would achieve the target exposure at steady state in a group of 100 virtual 3-month-olds. To achieve the same exposure as in Ayoub et al.^9^, a dose of 25 mg/kg metformin would be required.

### (C) Outcomes

#### Feasibility

Early phase drug trials in infants and young children are challenging for recruitment, retention, and compliance. Therefore, we will assess: (1) The total number of eligible patients during the study period; (2) The proportion of eligible patients that are approached for participation; (3) The proportion of approached patients that consent to trial inclusion; (4) The proportion of included patients that initiates the study medication, reaches the end of the study with less than 3 doses missed, or need premature discontinuation.

#### Safety

No serious complications have been reported with the use of metformin in the pediatric population. Lactic acidosis is the most severe complication and is usually observed in the context of overdose. Lactic acidosis may also occur with renal failure.^14^ Kidney and liver function will be assessed prior to the initiation of therapy, and with subsequent study visits. A change >2x from baseline will be considered an adverse event. While changes in serum glucose are unlikely, we will monitor for the incidence of hypoglycemia (glucose <60mg/dl) or hyperglycemia (glucose>140mg/dl). Mild gastrointestinal complaints, including vomiting or diarrhea, have been identified in pediatric and adult clinical trials of metformin. Therefore, we will ask parents/guardians to record feeding patterns (feed frequency and volume), the number of stools per day and any vomiting in the study diary. Severe gastrointestinal symptoms will be defined as >3 episodes of vomiting or >3 episodes of diarrhea per day; all other gastrointestinal symptoms will be classified as mild.

#### Stakeholder satisfaction

We will ascertain key facilitators and barriers that may impact engagement in future trials. Qualitative interview methods will be utilized to assess their experience with the recruitment process, study visits, and study tasks such as medication administration and diary keeping. Collecting this meaningful data will allow us to understand elements that impact enrollment and study compliance. A survey will be provided at study completion to elicit their feedback. An additional survey will be provided to parents/caregivers who decline to participate or withdraw from the study to understand the factors impacting these choices. All survey responses will be kept confidential.

#### Pharmacokinetics

Plasma metformin levels will be analyzed by investigators from blood obtained at study visits. Additionally, an optional sub-study of children with at-home monitoring will provide denser pharmacokinetic data between hospital-based study visits. Plasma metformin levels and time after metformin initiation will be added to the PBPK models described above. In keeping with what is done in industry for model confirmation during pediatric trials, the simulations, and therefore dose, will be considered appropriate when 90% of the observed data points fall within the 90th prediction interval of the population. If this is found to not be the case, sensitivity analysis will be completed to understand influential model parameters that can be updated, and doses in future trials will be refined.

### (D) Procedures

Infants enrolled in this study will complete a pre-study visit with baseline bloodwork including a complete blood count (CBC), liver and renal function, basic chemistry, glucose, and lactate. At this visit, parents will be taught how to administer metformin and given a 6-week supply of metformin at 50% of the target dose (12.5 mg/kg) to minimize potential gastrointestinal upset at higher doses. Parents will be educated on adverse effects and receive a diary to log dose administration and noted side effects. After six weeks, participants will return for a study visit with repeat labs. Bloodwork at this visit will include plasma metformin levels for measurement of pharmacokinetics. Study staff will also review diary completion for adverse effects and to ensure there have been no missed doses. Parents will then receive a 6-week supply of metformin 25mg/kg for 6 weeks. A final study visit will then occur following 12-weeks of metformin therapy, with repeat labs including plasma metformin levels. Participants will be deemed to have completed the study after the 12-week study visit. Of note, we will also enroll patients in an optional sub-study in which blood will be collected one week after initiation of low dose (12.5mg/kg) and target dose (25 mg/kg) metformin to measure plasma metformin levels and provide more granular pharmacokinetic data.

## ETHICS AND DISSEMINATION

Prior to trial commencement, approval of the Boston Children’s Hospital Institutional Review Board (IRB) will be obtained. Parents/caregivers of study participants will provide written informed consent. Trial oversight will be under the direction of a Data Safety Monitoring Board (DSMB) composed of individuals with the appropriate expertise, including external neonatologists and pharmacologists. Members of the DSMB will be independent from the study conduct and free of conflict of interest. The DSMB will meet at least semiannually to assess the safety of the study, with additional meetings invoked as required. The DMSB will operate under the rules of an approved charter that will be written and reviewed at the organizational meeting. Each data element the DSMB needs to assess will be clearly defined at the organizational meeting. The PI will be responsible for trial monitoring and verifying the rights and well-being of participants are protected, the reported trial data are accurate, complete, and verifiable from source documents, and that the conduct of the trial follows the currently approved protocol/amendments, local regulations and requirements, and guidelines from the International Council for Harmonisation of Technical Requirements for Pharmaceuticals for Human Use.

## DISCUSSION

There is an urgent need to establish new, innovative, adjunct therapies for HIE to shield infants against the sequalae of perinatal brain injury. While recent *in vitro* and *in vivo* studies have identified metformin as being potentially neuroprotective through benefits on neurogenesis and neuroplasticity, metformin has not been administered to infants after perinatal brain injury. Completion of this proposed trial will delineate the safety and feasibility of metformin use in infants after HIE. This study will also provide valuable pharmacokinetic data for metformin use in infants, addressing this research gap and informing a future phase II clinical trial on the effectiveness of the intervention.

Moreover, this trial provides a unique opportunity to partner with families and gather qualitative data regarding facilitators and barriers to study involvement and compliance, which will then be used to further inform a future phase II trial. This trial also presents opportunity to partner with non-profit organizations aimed at augmenting quality of life for families impacted by HIE through building awareness, advocacy, knowledge and support. Our plan for patient and community engagement will ensure timely communication and dissemination of study findings.

## CONCLUSION

This trial will demonstrate the safety and feasibility of metformin use in infants with a history of perinatal HIE, provide infant metformin pharmacokinetic data, and information regarding parent/caregiver satisfaction with the study processes and intervention. The data gathered in this trial will be used to inform and plan an efficacy-targeted clinical trial.

## Data Availability

All data produced in the present work are contained in the manuscript.

## AUTHORS CONTRIBUTIONS

All authors contributed to study design. The manuscript was primarily written by BTK and AH, with input from all authors. The pharmacokinetic analysis was performed by ANE.

## REFERENCES

1. Ferriero, D.M. (2004). Neonatal brain injury. N. Engl. J. Med. 351, 1985–1995. 10.1056/NEJMra041996.

2. Ma, T., Cg, W., Mj, V., Rk, W., and Da, S. (2012). Hypothermia for neonatal hypoxic ischemic encephalopathy: an updated systematic review and meta-analysis. Arch. Pediatr. Adolesc. Med. 166. 10.1001/archpediatrics.2011.1772.

3. Wu, Y.W., Comstock, B.A., Gonzalez, F.F., Mayock, D.E., Goodman, A.M., Maitre, N.L., Chang, T., Van Meurs, K.P., Lampland, A.L., Bendel-Stenzel, E., et al. (2022). Trial of Erythropoietin for Hypoxic-Ischemic Encephalopathy in Newborns. N. Engl. J. Med. 387, 148–159. 10.1056/NEJMoa2119660.

4. McAdams, R.M., and Berube, M.W. (2021). Emerging therapies and management for neonatal encephalopathy-controversies and current approaches. J. Perinatol. Off. J. Calif. Perinat. Assoc. 41, 661–674. 10.1038/s41372-021-01022-9.

5. Ruddy, R.M., Adams, K.V., and Morshead, C.M. (2019). Age- and sex-dependent effects of metformin on neural precursor cells and cognitive recovery in a model of neonatal stroke. Sci. Adv. 5, eaax1912. 10.1126/sciadv.aax1912.

6. Liu, Y., Tang, G., Zhang, Z., Wang, Y., and Yang, G.-Y. (2014). Metformin promotes focal angiogenesis and neurogenesis in mice following middle cerebral artery occlusion. Neurosci. Lett. 579, 46–51. 10.1016/j.neulet.2014.07.006.

7. Dadwal, P., Mahmud, N., Sinai, L., Azimi, A., Fatt, M., Wondisford, F.E., Miller, F.D., and Morshead, C.M. (2015). Activating Endogenous Neural Precursor Cells Using Metformin Leads to Neural Repair and Functional Recovery in a Model of Childhood Brain Injury. Stem Cell Rep. 5, 166–173. 10.1016/j.stemcr.2015.06.011.

8. Ameen, O., Samaka, R.M., and Abo-Elsoud, R.A.A. (2022). Metformin alleviates neurocognitive impairment in aging via activation of AMPK/BDNF/PI3K pathway. Sci. Rep. 12, 17084. 10.1038/s41598-022-20945-7.

9. Ayoub, R., Ruddy, R.M., Cox, E., Oyefiade, A., Derkach, D., Laughlin, S., Ades-Aron, B., Shirzadi, Z., Fieremans, E., MacIntosh, B.J., et al. (2020). Assessment of cognitive and neural recovery in survivors of pediatric brain tumors in a pilot clinical trial using metformin. Nat. Med. 26, 1285–1294. 10.1038/s41591-020-0985-2.

10. Flory, J., and Lipska, K. (2019). Metformin in 2019. JAMA 321, 1926–1927. 10.1001/jama.2019.3805.

11. Yeung, C.H.T., Verstegen, R.H.J., Greenberg, R., and Lewis, T.R. (2023). Pharmacokinetic and pharmacodynamic principles: unique considerations for optimal design of neonatal clinical trials. Front. Pediatr. 11, 1345969. 10.3389/fped.2023.1345969.

12. Hanke, N., Türk, D., Selzer, D., Ishiguro, N., Ebner, T., Wiebe, S., Müller, F., Stopfer, P., Nock, V., and Lehr, T. (2020). A Comprehensive Whole-Body Physiologically Based Pharmacokinetic Drug-Drug-Gene Interaction Model of Metformin and Cimetidine in Healthy Adults and Renally Impaired Individuals. Clin. Pharmacokinet. 59, 1419–1431. 10.1007/s40262-020-00896-w.

13. Sánchez-Infantes, D., Díaz, M., López-Bermejo, A., Marcos, M.V., de Zegher, F., and Ibáñez, L. (2011). Pharmacokinetics of metformin in girls aged 9 years. Clin. Pharmacokinet. 50, 735–738. 10.2165/11593970-000000000-00000.

14. Spiller, H.A., and Quadrani, D.A. (2004). Toxic effects from metformin exposure. Ann. Pharmacother. 38, 776–780. 10.1345/aph.1D468.

